# Clinical and attitudinal outcomes of a personalized, multimodal lifestyle intervention for Alzheimer’s disease prevention in high-risk, cognitively normal older adults in north Alabama: A pilot study

**DOI:** 10.64898/2026.09.19.26363495

**Authors:** Meagan Cochran, Jada Pugh, Tanner Coleman, Whitley Kelley, Paige Jamieson, Elizabeth Dunne, Patti Smith, Jessica Chasteen, Mary Jones, Jenny Turner, Halle Burns, Gwendolyn Geissler, Mary Brunkow, S. Quinn Johnston, Katherine Metzger, Lena Seyfarth, Jared Taylor, Emily Shields, William Herrington, Wayne Schroer, Scott Newberry, Anna C.E. Hurst, James Yurkovich, Neil Lamb, Lee Hood, Simon Evans, Florencia P. Behn, Jennifer Lovejoy, J. Nicholas Cochran

## Abstract

**INTRODUCTION:** Multimodal lifestyle interventions can delay or slow Alzheimer’s disease (AD) in at-risk older adults, and plasma pTau217, a blood-based biomarker, now enables scalable AD risk stratification. Population-level screening integrating genetic, biomarker, and lifestyle risk factors, alongside attitudes toward risk disclosure, remains understudied. This pilot assessed dementia-risk attitudes before and after disclosure and tested a personalized, data-driven multimodal coaching intervention in a high-risk Alabama population.

**METHODS:** Cognitively normal adults aged 65–75 with a first-degree relative affected by AD/dementia were recruited in north Alabama and screened using the Montreal Cognitive Assessment, family history, *APOE* genotyping, non-*APOE* AD polygenic risk score, and plasma pTau217. A composite rubric classified high-risk status (score >5 and/or pTau217 >0.18 pg/mL); high-risk participants received results disclosure and a 6-month personalized intervention (dietitian-led coaching, cognitive training, activity tracking). Pre– and post-disclosure attitudes, distress (REVEAL IGT-AD scale), and biomarker/cognitive measures were assessed.

**RESULTS:** Of 138 screened participants, 52 (37.7%) were high-risk, most (69.2%) by pTau217 alone. Before disclosure, subjective dementia-risk perception correlated with objective risk score. Scores consistent with post-disclosure clinically significant distress occurred in 28% of survey respondents in at least one timepoint but was often (50%) observed with pre-existing anxiety/depression. Over 6 months, MoCA scores and lifestyle risk indices were unchanged, pTau217 continued rising but at a slowed rate compared to between screening and the beginning of the lifestyle intervention, and self-reported worry and perceived dementia risk decreased significantly.

**DISCUSSION:** A genomics– and biomarker-informed screening and coaching model is feasible in a high-risk cohort in the deep south, with manageable distress and improved attitudes despite limited short-term biological change. Findings support larger, longer, and more diverse trials validating this approach for AD screening.

## Introduction

A number of studies have pointed to the effectiveness of multimodal intensive lifestyle interventions (ILIs) for preventing or delaying late-onset Alzheimer’s disease (AD) when implemented in cognitively healthy older adults or those with early stage cognitive decline such as the FINGER study (Ngandu et al. 2015), the US POINTER study (Baker et al. 2025), and the LATAM-Fingers study (Crivelli et al. 2026). Multimodal ILIs, which typically include dietary modification, physical exercise, cognitive training, and social engagement, have previously been shown to be effective in presymptomic adults both with and without increased genetic risk for AD, i.e. *APOE* ε4 alleles. While most studies have implemented a standardized ILI to all participants, the COCOA study (Coaching for Cognition in Alzheimer’s Disease study) demonstrated that a data-driven multimodal health coaching intervention, leveraging both genetic and broad clinical data to personalize interventions, significantly improved both cognitive and functional scores in patients with early stage AD (Roach et al. 2022, Roach et al. 2023).

An important development in recent years has been the assessment of peripheral blood biomarkers for AD risk. Of these biomarkers, pTau217 has emerged as the most tractable, reliable, and reproducible biomarker indicative of the future development of AD pathology and disease (Schindler et al. 2024). Other studies have also tied defined risk levels to a variety of clinical measurements, including blood lipids (Solomon et al. 2009), hemoglobin A1c (Crane et al. 2013), blood pressure (Gottesman et al. 2017), and obesity (Whitmer et al. 2005). Given this, we now have more knowledge than ever to facilitate the prediction of dementia risk in the population and effective intervention options for those at high risk of conversion to AD. However, the field has yet to broadly implement population screening or gauge attitudes around risk before and after enlightening the general population about multimodal lifestyle interventions that may shift risk.

The purpose of this pilot study was two-fold. First, we sought to assess attitudes around dementia risk for participants as they learn of their personalized risk and possible lifestyle factors that may modify that risk. Secondly, we sought to implement a data-driven, personalized multi-modal coaching intervention in a rural population in north Alabama to see if the results of the COCOA study, which used a commercial wellness program, could be replicated.

## Methods

### Recruitment

Prospective participants were recruited via community events, social media, local news sources, and word of mouth from local healthcare providers. In order to be eligible for screening, individuals were required to be between the ages of 65 and 75 years old, have at least one first degree relative who has or had AD or related dementias, and self-identify as having no current cognitive impairment.

### Screening

Qualified individuals were enrolled at the Smith Family Clinic for Genomic Medicine in Huntsville, AL by certified genetic counselors. The Montreal Cognitive Assessment (MoCA 8.1) was administered by trained personnel, and a score of >25 was required for continuation in the study. Additional screening measures included family history collection, surveys regarding medical history, nutrition, exercise, cognitive activity, and attitudes regarding dementia risk, pTau217 (Labcorp, Inc; Burlington, NC) and *APOE* genotyping and AD polygenic risk score (PRS) testing (Allelica, Inc; New York, NY). Risk points were assigned based on a rubric developed for the study (**Table 1**).

**Table 1:** Points-based risk rubric to identify pre-symptomatic individuals at high risk for Alzheimer’s disease. The possible risk score range is −4 to 17. FDR: First Degree Relative. SDR: Second Degree Relative. mCAIDE: modified Cardiovascular Risk Factors, Aging, and Incidence of Dementia. MoCA: Montreal Cognitive Assessment. HF: Healthy factors.

| Points Assigned: | -2 | -1 | 0 | 1 | 2 | 3 |
| --- | --- | --- | --- | --- | --- | --- |
| **Family History** |  |  | No FDR | 1 FDR | More than Category 1, less than Category 3 | 2 FDR OR<br>1 FDR <65 OR<br>1 FDR + 2 SDR same side) |
| **APOE Genotype** | 2/2 | 2/3 | 3/3 | 2/4 OR<br>3/4 |  | 4/4 |
| **Non-APOE AD PRS** |  |  | Lower 85% | Top 15% |  |  |
| **mCAIDE Score** |  | 0-2 | 3-6 | 7-10 | 11-14 |  |
| **(Dhana et al. 2020) Score** |  | 4-5 HF | 3 HF | 2 HF | 0-1 HF |  |
| **MoCA** |  |  | 29-30 | 27-28 | 25-26 |  |
| **pTau217 (pg/mL)** |  |  | <0.18 | 0.18-0.25 | >0.25-0.32 |  |
| **Self-identified Black/AA or Hispanic** |  |  | No | Hispanic | Black/AA |  |

A total risk score of >5 and/or a positive pTau217 result (>0.18pg/mL) qualified for inclusion in the “high risk” cohort. Participants who did not qualify for the high risk cohort had their clinical results (pTau217, *APOE*, and PRS) returned via a personalized mailed result letter. Individuals in the high risk cohort returned to clinic for results disclosure by certified genetic counselors.

### Personalized Multimodal ILI

At the time of results disclosure, high risk individuals were invited to participate in a 6-month ILI consisting of personalized health coaching by a registered dietician, brain training via BrainHQ, and sleep and activity tracking via FitBit.

The content of the ILI was based on previous studies of lifestyle interventions for the prevention or delay of dementia, such as the Finnish Geriatric Intervention Study to Prevent Cognitive Impairment and Disability (FINGER) (Kivipelto et al. 2013, Ngandu et al. 2015), the Coaching for Cognition in Alzheimer’s (COCOA) trial (Roach et al. 2022, Roach et al. 2023), and the study by (Ornish et al. 2024). The intervention supported adoption and maintenance of healthy behaviors including healthy nutrition (MIND Diet), physical activity (150 minutes/week of moderate activity), stress management, weight management, and adherence to physician-prescribed medical regimens. In addition, participants were encouraged to participate regularly in social and learning activities, and to engage in cognitive training via Posit’s BrainHQ web-based training tools (brainhq.com). All information to support healthy behavior was based on standard, published, and well-researched health data. Participants were referred to their primary healthcare provider for any medical conditions or clinical risk factors discovered from the data.

Participants in the ILI were supported by telephone-based health coaching. In this pilot study, all coaching was provided by a single coach, who was a licensed Registered Dietitian Nutritionist. Coaching calls were scheduled monthly, with email or text communication available between the monthly calls. Participants could contact their coach at any time with questions or for support. Using the combination of an individual’s biological data (including genomics and clinical and nutritional assays), activity data, behavioral data, and other incoming streams of health information, the coach tailored the lifestyle intervention for each participant. The coach had access to participants’ BrainHQ data to encourage compliance with the recommended training regimen.

### Trial Measures

Upon initiation and conclusion of the lifestyle intervention trial, participants completed the following: MoCA 8.2 (at enrollment), MoCA 8.3 (at conclusion), pTau217, and clinical labs including heavy metals profile, CBC with differential, CMP, lipid panel, hemoglobin A1C, serum folate, TSH, vitamin D, Lp(a), CRP, serum MMA, homocysteine, uric acid, GGT, insulin, and ferritin. In addition, they completed surveys regarding their current health status, nutrition, sleep, exercise, cognitive activity, and attitudes regarding dementia risk. In addition, post-result distress (Risk Evaluation and Education for Alzheimer’s Disease Impact of Genetics Testing Distress Subscale) and measures of depression and anxiety (Geriatric Anxiety Scale and Geriatric Depression Scale) were completed post-disclosure and at the conclusion of the 6-month trial.

### Oversight and registration

This project was approved by WCG IRB (#20250931) and registered on ClinicalTrials.gov (NCT07146412).

### Statistics

Statistical tests are noted through the results and were calculated using GraphPad Prism 11.1.0.

## Results

### Enrollment

165 individuals sought screening; 6 declined enrollment after informed consent, 21 failed initial MoCA and were thus unable to continue, and 138 completed screening. 52 individuals were identified as high risk and proceeded to the 6-month trial. Of these, 69.2% (n=36) qualified by positive pTau217 alone, 9.6% (n=5) qualified by positive pTau217 and by risk points, and 21.2% (n=11) qualified by risk points alone.

### Baseline demographics

Both total and high-risk cohorts were predominantly female, white, and highly educated. Key summary baseline characteristics of the total cohort screened and the high-risk cohort enrolled in the intervention (as well as the subset of those not meeting high-risk criteria) is described in **Table 2**.

**Table 2:** Baseline characteristics of participants. No statistically significant differences between the total cohort and either high-risk or not high-risk subgroups were detected using the tests noted below.

| | Total cohort<br>(N=138) | High-risk<br>(N=52) | Not high-risk (N=86) | High / Not-high $\Delta$? |
| --- | --- | --- | --- | --- |
| Female (%) | 69 | 62 | 73 | ns by Fisher's exact |
| Self-reported race: white (%) | 93 | 90 | 94 | ns by Fisher's exact |
| Self-reported non-Hispanic (%) | 96 | 94 | 97 | ns by Fisher's exact |
| Mean age (years) | 68.8 | 69.4 | 68.4 | *$p=0.038$ by Mann-Whitney |
| Education: bachelors+ (%) | 84 | 77 | 90 | ns by Fisher's exact |
| Mean BMI (kg/m<sup>2</sup>) | 27.7 | 27.8 | 27.6 | ns by Mann-Whitney |
| Systolic BP (mm Hg) | 127.6 | 128.3 | 127.2 | ns by Mann-Whitney |

### Risk determinants and overall risk scores

In addition to traditionally measured baseline demographics, we collected study-specific information used to calculate the risk score summarized by **Table 1**. Key descriptive statistics of determinants of this risk score are noted in **Table 3**.

**Table 3:** Summary statistics for determinants of risk score described in **Table 1**. * in High-risk and Not high-risk columns indicates a significant difference from the total cohort by the test noted in the rightmost column where High-risk and Not high-risk are compared. ^#^Note that two measurements were missing for baseline pTau217 measures, so N=136 for Total cohort, 51 for High-risk, and 85 for Not high-risk for that measure. The High risk participant qualified based on factors outside of their baseline pTau217 measure.

| | Total cohort<br>(N=138)<sup>#</sup> | High-risk<br>(N=52)<sup>#</sup> | Not high-risk<br>(N=86)<sup>#</sup> | High-risk / Not high-risk $\Delta$? |
| --- | --- | --- | --- | --- |
| Family history (% strong / moderate / minimal) | 39/37/24 | 50/29/21 | 30/40/30 | *$p=0.039$ by Cochran-Armitage |
| Mean APOE Genotype Score | 0.33 | 0.69* | 0.10* | *$p<0.0001$ by Mann-Whitney |
| Mean Non-APOE AD PRS | 50.0 | 58.8 | 44.7 | *$p=0.0046$ by Mann-Whitney |
| Mean mCAIDE Score | 4.5 | 5.1 | 4.2 | *$p=0.0130$ by Mann-Whitney |
| (Dhana et al. 2020) Score | 3.9 | 3.6 | 4.1 | *$p=0.0045$ by Mann-Whitney |
| MoCA | 27.6 | 27.1 | 27.9 | *$p=0.0038$ by Mann-Whitney |
| pTau217 (pg/mL) | 0.18 | 0.29* | 0.11* | *$p<0.0001$ by Mann-Whitney |
| Self-identified Black/AA or Hispanic Score (0/1/2) | 127/6/5 | 45/3/4 | 82/3/1 | *$p=0.037$ by Cochran-Armitage |
| Mean non-pTau Table 1 score | 3.0 | 4.4* | 2.2* | *$p<0.0001$ by Mann-Whitney |

### Attitudes towards dementia risk before results disclosure

In addition to biological measures, we surveyed participants at screening, before they learned of their risk status. Those results are summarized by **Table 4**, noting that the categories reflect the category that the participant ultimately qualified for, but that these survey results preceded their knowledge of the category.

**Table 4:**
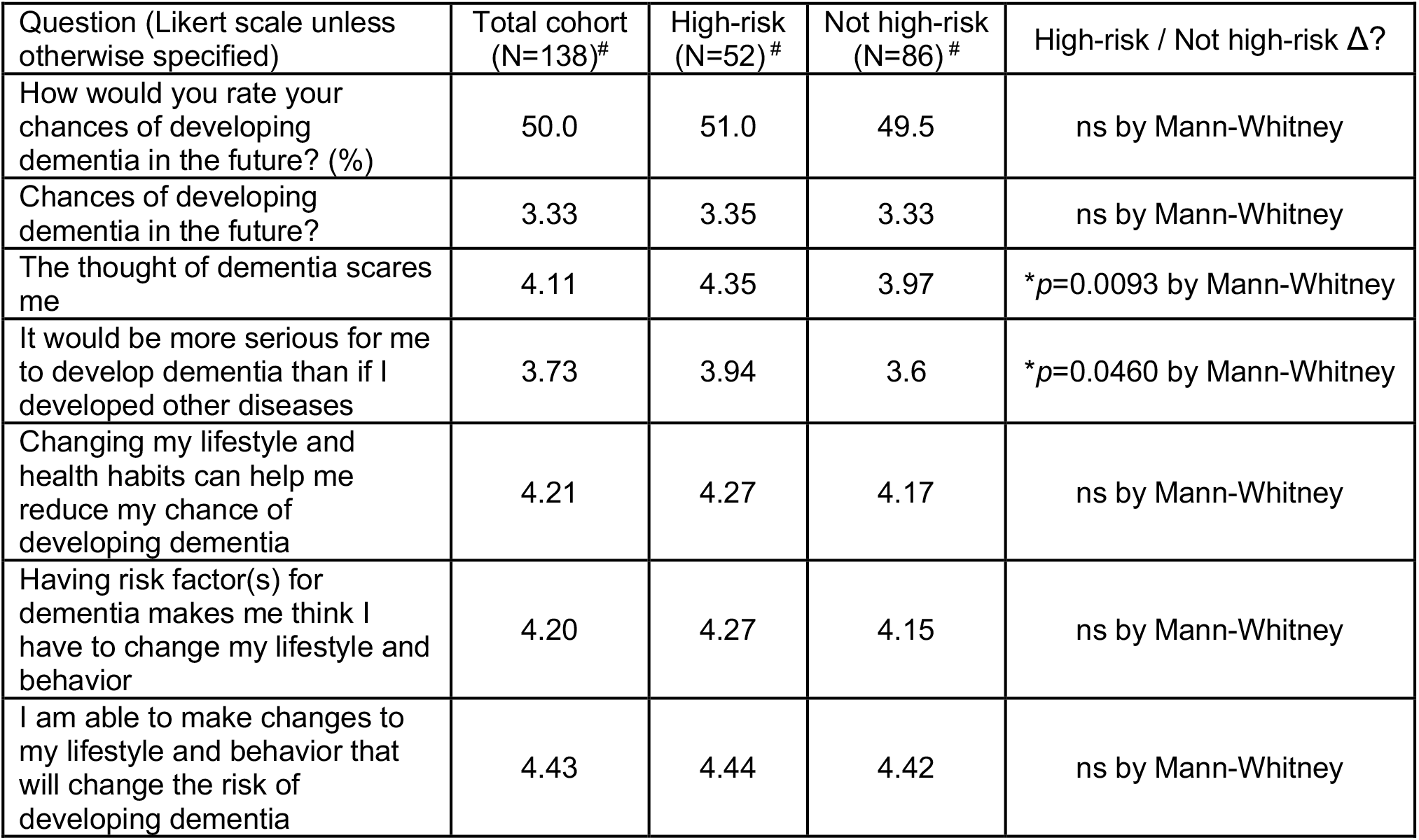
Participant attitudes towards learning of dementia risk prior to results disclosure (but presented by group that they ultimately qualified for).

| Question (Likert scale unless otherwise specified) | Total cohort<br>(N=138)<sup>#</sup> | High-risk<br>(N=52)<sup>#</sup> | Not high-risk<br>(N=86)<sup>#</sup> | High-risk / Not high-risk $\Delta$? |
| --- | --- | --- | --- | --- |
| How would you rate your chances of developing dementia in the future? (%) | 50.0 | 51.0 | 49.5 | ns by Mann-Whitney |
| Chances of developing dementia in the future? | 3.33 | 3.35 | 3.33 | ns by Mann-Whitney |
| The thought of dementia scares me | 4.11 | 4.35 | 3.97 | *$p=0.0093$ by Mann-Whitney |
| It would be more serious for me to develop dementia than if I developed other diseases | 3.73 | 3.94 | 3.6 | *$p=0.0460$ by Mann-Whitney |
| Changing my lifestyle and health habits can help me reduce my chance of developing dementia | 4.21 | 4.27 | 4.17 | ns by Mann-Whitney |
| Having risk factor(s) for dementia makes me think I have to change my lifestyle and behavior | 4.20 | 4.27 | 4.15 | ns by Mann-Whitney |
| I am able to make changes to my lifestyle and behavior that will change the risk of developing dementia | 4.43 | 4.44 | 4.42 | ns by Mann-Whitney |

While the question “How would you rate your chances of developing dementia in the future?” did not differ between groups, it did exhibit an overall positive correlation with the aggregate risk score inclusive of pTau217 (Spearman \**p*=0.0378 for numerical, \**p*=0.0256 for Likert) as well as excluding pTau217 (Spearman \**p*=0.0282 for numerical, \**p*=0.0108 for Likert). “The thought of dementia scares me” was also positively correlated with risk score inclusive of pTau217 (Spearman \**p*=0.0465).

### Post-disclosure distress results for high-risk participants

In addition to assessing attitudes towards dementia risk before results disclosure, we also evaluated distress after disclosure using the Risk Evaluation and Education for Alzheimer’s Disease Impact of Genetics Testing Distress (REVEAL IGT-AD) scale (Chung et al. 2009). A recent study has suggested a threshold of >15 in the context of disclosure of amyloid PET positivity disclosure (Ren et al. 2025) for recommendation of additional psychological support. Of the 52 participants enrolled in the intervention, 35 completed post-disclosure distress surveys. Of those completed, 10 (29% of respondents and 19% of total participants) had REVEAL IGT-AD scores >15. We also re-evaluated distress at 6 months post disclosure. Because these surveys were conducted in-clinic rather than remotely, there was a higher response rate of 48 participants, out of which 10 (6 common between disclosure and 6 months, 21% of respondents and 19% of total participants) had REVEAL IGT-AD scores >15. Out of 14 participants who had a score of >15 at either time point, 7/14 (50%) had diagnoses preceding this study of depression and/or anxiety compared to 9/36 (25%) of those who responded to at least one survey but had scores <15, but this difference was not significant by Fisher’s exact test (*p*=0.11). Of those who completed at least 1 survey, 36/50 (72%) had scores <15, 8 (16%) had scores >15 at 1 timepoint, and 6 had scores >15 at both timepoints.

### Intervention results

The goal of this pilot study was not to evaluate the effectiveness of the ILI, but rather to conduct a logistical pilot and to gauge participant attitudes and distress to receiving high risk information. Nevertheless, here we present key exploratory results pre– and post-intervention, summarized in **Table 5**.

**Table 5:** Intervention results. ^#^Note that two measurements were missing for pTau217, 1 at screening and 1 at return of results, so N=51 for each of those timepoints for pTau217. Statistical comparisons were Wilcoxon matched-pairs signed rank test.

|  | Screening<br>(average -3<br>months)<br>(N=52) | Return of<br>Results<br>(0 months)<br>(N=52) | Post-Intervention<br>(6 months)<br>(N=50) | Screening<br>vs. RoR | Post-Intervention<br>vs. RoR | Post-Intervention<br>vs.<br>Screening |
| --- | --- | --- | --- | --- | --- | --- |
| Average MoCA | 27.1 | 27.0 | 26.7 | ns | ns | ns |
| Average pTau217 (pg/mL) | 0.270 | 0.295 | 0.311 | *$p=0.042$ | ns, $p=0.228$ | *$p=0.0002$ |
| Average Dhana score | 3.56 | 3.58 | 3.65 | ns | ns | ns |
| Average mCAIDE score | 5.08 | 5.23 | 5.16 | ns | ns | ns |
| Chance of future dementia<br>self-report % | 51.0 | 50.8 | 45.5 | ns | *$p=0.0293$ | ns |
| Thought of dementia<br>Scares Me | 4.35 | 4.35 | 3.96 | ns | *$p=0.0103$ | *$p=0.0150$ |

While lifestyle scores did not detectably change over this short 6 month intervention period, there was a notable non-zero slope via linear regression when comparing the change in Dhana lifestyle score and the change in MoCA Post-Intervention vs. RoR (*p=0.02 by F test, **Figure 1**).

**Figure 1:**
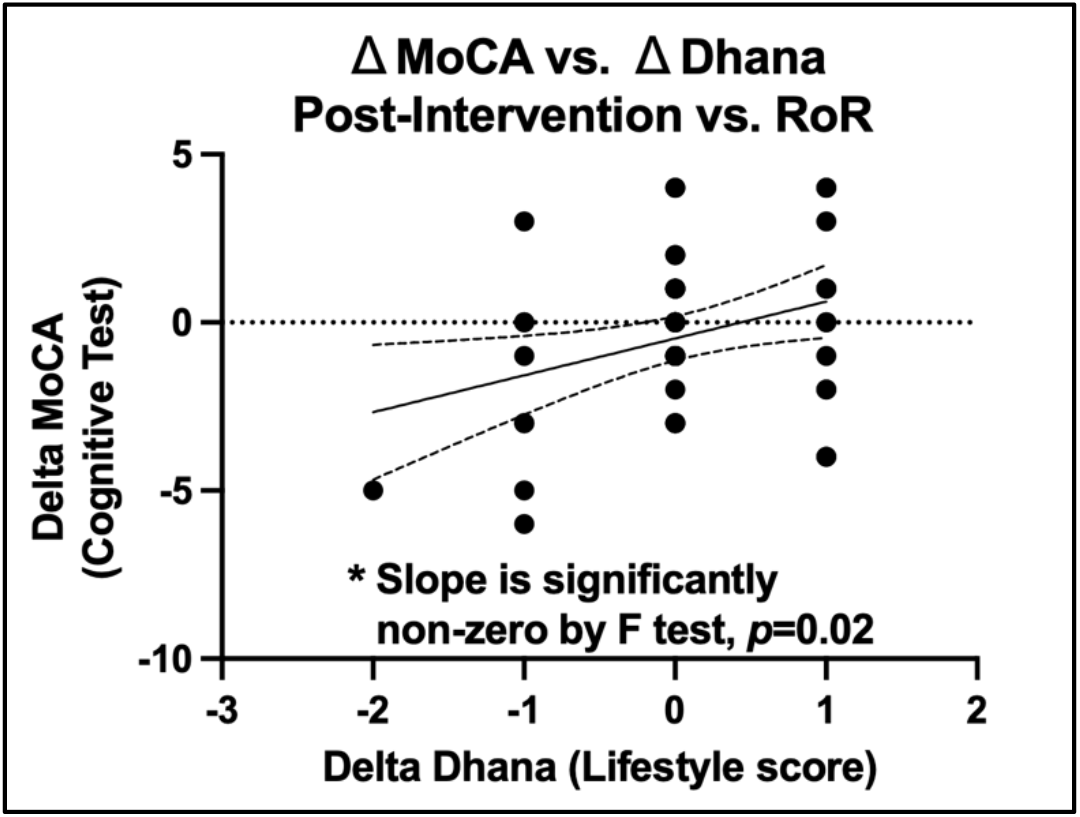
Linear regression of the change in MoCA vs. the change in Dhana score comparing Post-intervention (6 month) to Return of results (0 months). Slope is 1.097, and is significantly non-zero by F test (\**p*=0.0205).

## Discussion

This pilot study demonstrates the feasibility of a genomically– and biomarker-informed approach to population-level Alzheimer’s disease (AD) risk stratification, coupled with a personalized, data-driven lifestyle coaching intervention, in a community-recruited population in north Alabama. Among 138 cognitively normal older adults with a first-degree relative affected by AD or a related dementia, more than one-third (37.7%, n=52) met criteria for a composite “high-risk” designation using a rubric that integrated family history, *APOE* genotype, a non-*APOE* AD polygenic risk score, cardiovascular and lifestyle risk indices, cognitive screening, plasma pTau217, and self-identified race/ethnicity. A positive pTau217 result alone accounted for the majority (69.2%) of high-risk classifications, reinforcing the growing consensus that this blood-based biomarker is sufficiently sensitive, reproducible, and scalable to serve as an anchor for population-level AD risk screening outside of specialized memory clinics (Palmqvist et al. 2024, Schindler et al. 2024). Notably, before learning any individual results, participants’ subjective sense of their own dementia risk already correlated with their eventual objective risk score, suggesting that adults with a family history of dementia carry meaningful, if imprecise, insight into their own risk even before formal testing.

The results of the REVEAL IGT-AD scale are particularly important to highlight in this study as this is the first study (to our knowledge) to apply this paradigm to pre-symptomatic return of multimodally-determined risk using a combination of *APOE*, non-*APOE* AD PRS, and pTau217 biomarker results. 14/50 (28%) of high risk participants had scores consistent with clinically significant distress in at least 1 time point, but 50% of these individuals had pre-existing diagnoses of anxiety and/or depression. Importantly, 36/50 (72%) of participants who completed at least one survey never had a score consistent with clinical significant distress. Our observed rates are comparable to distress rates reported after amyloid PET and *APOE* genotype disclosure in other cognitively unimpaired cohorts (Ketchum et al. 2024, Grill et al. 2026), and is consistent with two decades of disclosure research beginning with the original REVEAL trial of *APOE* genotype disclosure, which similarly found no significant excess of short-term anxiety or depression attributable to disclosure itself, with pre-existing emotional vulnerability being the strongest predictor of post-disclosure distress (Green et al. 2009). Our finding that a numerically (though not statistically significantly) higher proportion of highly distressed participants had a pre-existing diagnosis of depression or anxiety is consistent with this pattern and suggests that brief pre-disclosure psychological screening, rather than withholding risk information altogether, may be the more productive strategy as this type of program scales.

The exploratory intervention data should be interpreted cautiously, as this pilot was neither designed nor powered to test the efficacy of the multimodal lifestyle intervention (ILI). Over the 6-month coaching period, objective measures of cognition (MoCA) and composite lifestyle risk (Dhana score, mCAIDE score) did not change significantly, and pTau217 continued to rise, consistent with the expected trajectory of this biomarker over time and the fact that six months is brief relative to the years-long timescale over which AD blood biomarkers accumulate (Ossenkoppele et al. 2022) and over which prior multidomain trials have demonstrated benefit (Ngandu et al. 2015, Baker et al. 2025, Crivelli et al. 2026). Still, over the course of this pilot, ptau217 levels increased slightly but significantly from screening to initial return of results (an average of 79 days). However, while pTau217 levels significantly increased from screening to the final timepoint (an average of 257 days), they did not detectably statistically change during the course of the 6 month intervention (an average of 177 days). This raises the intriguing speculation that the ILI may have slowed the rate of pTau217 increase; indeed, by averages, the rate of increase slowed from 0.32 femtograms/day to 0.09 femtograms/day, a 3.6 fold effect.

Despite the absence of or speculative nature of changes in ILI-related measures, participants’ self-reported worry about developing dementia and their perceived likelihood of future dementia both decreased significantly over the same period. This dissociation between subjective and objective outcomes may reflect a therapeutic or “empowerment” effect of sustained personalized coaching contact itself, independent of measurable biological or cognitive change, a pattern that echoes the psychosocial benefits described in the COCOA trial, on which our intervention content was modeled (Roach et al. 2022, Roach et al. 2023). We also observed a nominally significant, hypothesis-generating association between the magnitude of an individual’s improvement in lifestyle risk score and their change in MoCA performance, which, while underpowered to draw firm conclusions, is directionally consistent with the dose-dependent relationships between lifestyle modification and cognitive outcomes reported in FINGER, US POINTER, and LatAm-FINGERS (Ngandu et al. 2015, Baker et al. 2025, Crivelli et al. 2026).

### Limitations

An important limitation of this pilot, with direct relevance to its goal of informing population screening in Alabama, is the lack of racial and ethnic diversity in both the screened and high-risk cohorts (93% white, 96% non-Hispanic), despite Black and African American individuals experiencing a disproportionately higher incidence of AD nationally (Barnes 2022) and higher proportion of Black and African American individuals in Alabama (~26%) than the recruited population in the cohort. Our risk rubric deliberately incorporated additional points for self-identified Black/African American or Hispanic ethnicity in an effort to increase sensitivity for populations known to be under-ascertained by standard risk tools, but this design choice alone was not sufficient to overcome the recruitment and access barriers that limited enrollment of these groups in a community-based, word-of-mouth recruitment model. Given that a substantial proportion of dementia risk is attributable to potentially modifiable factors (Livingston et al. 2024), and that Alabama’s population includes a substantial proportion of Black residents who bear a disproportionate burden of AD and its vascular risk factors inclusive of many of these modifiable risk factors, achieving equitable representation will be essential for any population-level screening and intervention program, including the broader initiative this pilot is intended to inform.

Beyond the demographic homogeneity described above, this study has several additional limitations. First, and most fundamentally, it was a single-arm, pre-post pilot without a concurrent control or comparison group; consequently, we cannot attribute the observed reductions in dementia-related worry, or the absence of change in objective measures, specifically to the content of the ILI as opposed to regression to the mean, practice effects, or the non-specific benefits of sustained personalized contact with a coach and study team. Second, the sample was small (52 high-risk participants) and recruited from a single region via community outreach, word of mouth, and self-referral, introducing volunteer bias toward individuals who were already health-engaged and limiting generalizability to the broader population that an eventual screening program would need to serve. Third, the 6-month intervention window is short relative to the multi-year timescales used in comparable multidomain trials (2 years in FINGER and US POINTER) and is unlikely to be sufficient to detect meaningful change in slowly progressive biomarkers such as pTau217 or in hard cognitive endpoints. Fourth, all coaching in this pilot was delivered by a single registered dietitian nutritionist; while this maximized consistency, it leaves open the question of whether outcomes are generalizable across coaches and scalable to a larger, statewide program without loss of fidelity. Fifth, lifestyle, nutrition, sleep, and exercise data relied heavily on self-report surveys, which are subject to recall and social desirability bias, although objective FitBit and BrainHQ engagement data were also collected and may be examined in future analyses. Sixth, survey response rates varied across timepoints and modality (for example, 35 of 52 for remote post-disclosure distress surveys versus 48 of 52 for in-clinic 6-month surveys), raising the possibility of non-response bias in the distress estimates. Seventh, statistical comparisons are nominal and are not adjusted for multiple testing, and several results were only marginally significant (p-values near 0.05); these findings should therefore be considered exploratory and hypothesis-generating rather than confirmatory. Finally, the >15 distress threshold applied to the REVEAL IGT-AD Distress subscale was empirically derived and validated in the context of amyloid PET disclosure in individuals with MCI or dementia (Ren et al. 2025), not in the context of combined genetic and blood-based biomarker disclosure to cognitively normal adults as used here; its direct applicability to our disclosure paradigm is a reasonable but unconfirmed assumption.

### Future directions

These findings support several concrete next steps. Foremost, a larger, adequately powered trial with a randomized or delayed-start control arm and a longer follow-up period (ideally 18–24 months, in line with FINGER, US POINTER, and LatAm-FINGERS) is needed to determine whether this personalized, biomarker– and genetics-informed coaching model measurably alters cognitive trajectories or biomarker slopes relative to education or standard care alone. Second, future recruitment efforts must proactively engage Black and African American, Hispanic, and other underrepresented communities in Alabama through community health workers, faith-based organizations, and federally qualified health centers, both to validate the risk rubric in a more representative population and to ensure that any resulting population screening program reduces, rather than widens, existing disparities in AD diagnosis and care (Barnes 2022). Third, the composite risk rubric itself warrants prospective validation against hard outcomes (incident MCI or dementia, longitudinal biomarker trajectories) in a larger and more diverse cohort to refine its component weightings and thresholds. Fourth, the hypothesis-generating association between lifestyle score improvement and cognitive change should be explored mechanistically using the objective BrainHQ and FitBit adherence data already collected to identify which intervention components (diet, physical activity, cognitive training, coaching contact frequency) are most strongly associated with benefit. Fifth, given the relatively low but non-trivial rate of clinically significant post-disclosure distress observed here and elsewhere (Ketchum et al. 2024, Grill et al. 2026), future work should evaluate brief pre-disclosure psychological screening to identify participants who may benefit from additional support and should formally validate distress instruments such as the IGT-AD for use in combined genetic-andblood-biomarker disclosure contexts. Finally, as plasma pTau217 assays become more widely available in primary care settings (Palmqvist et al. 2024), health-economic and implementation-science evaluations will be needed to determine how a coaching-supported population screening model of this kind can be scaled cost-effectively and sustainably across Alabama, including through lower-cost delivery formats such as group coaching, digital/telehealth platforms, and remote screening paradigms (Huber et al. 2024, Huber et al. 2026, Schultz et al. 2026).

## Data Availability

All data produced in the present study are available upon reasonable request to the authors.

## Acknowledgements

We thank Eric Verdin, Remy Gross, Lisa Ellerby, Brianna Stubbs, Birgit Schilling, and Simon Melov at the Buck Instiute for Research on Aging, Jeff Boore from Phenome Health, Cory Funk from Fulcrum and the Institute for Systems Biology, and Brian Pollock, Richard Myers, Danny Windham, and Carter Wells from HudsonAlpha for participation in discussions on study design.

